# Optimizing one-dose and two-dose cholera vaccine allocation in outbreak settings: A modeling study

**DOI:** 10.1101/2021.11.18.21266550

**Authors:** Tiffany Leung, Julia Eaton, Laura Matrajt

**Affiliations:** Vaccine and Infectious Disease Division, Fred Hutchinson Cancer Research Center, Seattle, WA, USA; School of Interdisciplinary Arts and Sciences, University of Washington, Tacoma, WA, USA

**Keywords:** cholera, vaccine optimization, mathematical modeling

## Abstract

**Background:** A global stockpile of oral cholera vaccine (OCV) was established in 2013 for use in outbreak response and are licensed as two-dose regimens. Vaccine availability, however, remains limited. Previous studies have found that a single dose of OCV may provide substantial protection against cholera.

**Methods:** Using a mathematical model with two age groups paired with optimization algorithms, we determine the optimal vaccination strategy with one and two doses of vaccine to minimize cumulative overall infections, symptomatic infections, and deaths. We explore counterfactual vaccination scenarios in three distinct settings: Maela, the largest refugee camp in Thailand, with high in- and out-migration; N’Djamena, Chad, a densely populated region; and Haiti, where departments are connected by rivers and roads.

**Results:** Over the short term under limited vaccine supply, the optimal strategies for all objectives prioritize one dose to the older age group (over five years old), irrespective of setting and level of vaccination coverage. As more vaccine becomes available, it is optimal to administer a second dose for long-term protection. With enough vaccine to cover the whole population with one dose, the optimal strategies can avert up to 30% to 90% of deaths and 36% to 92% of symptomatic infections across the three settings over one year. The one-dose optimal strategies can avert 1.2 to 1.8 times as many cases and deaths as a two-dose pro-rata strategy.

**Conclusions:** In an outbreak setting, speedy vaccination campaigns with a single dose of OCV may avert more cases and deaths than a two-dose pro-rata campaign under a limited vaccine supply.

## 1 Introduction

Cholera is an infectious disease caused by *Vibrio cholerae* with an estimated 1.3 to 4 million cases and 21,000 to 143,000 deaths annually [1], particularly in sub-Saharan Africa or displaced populations. Improved access to water, sanitation, and hygiene (WASH) is the mainstay of long-term cholera control and prevention. For the short to medium term, until WASH improvements are established, vaccination can serve as a key strategy in cholera control and prevention.

In 2013, the World Health Organization (WHO) created a global stockpile of oral cholera vaccine (OCV) to ensure access to OCVs in outbreak and humanitarian emergency situations [2]. OCV is administered in two doses, given two weeks apart, and is much less effective in young children than in adults. Vaccine efficacy for two doses is 42% in those under five years old and 72% in those five years and older against clinical cholera [3]. For one dose, vaccine efficacy is reduced to 8% for those under five years old and 57.5% for those five years and older [4, 5]. However, protection from the vaccine wanes over time. Children under five years old bear the greatest burden of cholera [6].

The standard approach to mass vaccination campaigns of OCV is untargeted and administers two doses [7, 8, 9, 10]. There remains, however, a global shortage of OCV. Countries have explored ways to maximize impact of limited vaccine supply. In April 2016 during the early stages of a cholera outbreak in Zambia, the Ministry of Health, in collaboration with Médicins sans Fronti`eres (MSF) and the WHO, implemented a single-dose campaign as a way to stretch vaccine supply [11, 12]. Eight months later in December, a second dose was administered when there was more stock [12]. A single-dose approach was also used for cholera outbreaks in South Sudan in 2015 and was effective at preventing medically attended cholera during an outbreak [13]. A one-dose OCV campaign would vaccinate more people in exchange for a lower level of protection.

The optimal use of vaccine depends on many factors, such as the objectives of a vaccination program, the number of doses available, vaccine efficacy and a region’s local transmission dynamics. Local transmission dynamics may be influenced by region-specific patterns in migration, human mobility, and climate variability [14, 15, 16, 17, 18].

In this study, we use mathematical models paired with optimization algorithms to determine the optimal vaccine allocation strategies. We minimized three metrics of infection and disease burden: cumulative infections, cumulative symptomatic infections, and cumulative deaths. We explore the impact OCV could have had in outbreaks in Chad, Thailand, and Haiti—three distinct settings representing an urban area, a refugee camp, and a region torn by natural disaster respectively. For these settings each with distinct local transmission dynamics, minimizing each metric produced a different optimal vaccination strategy. We quantify the impact of these optimal strategies through these metrics.

## 2 Methods

Here we describe the methods we used for the study. We provide details of each outbreak setting. We describe the three mathematical models used per setting and highlight the distinctive features of each. Next, we elaborate on the implementation of the vaccination campaign and the various vaccination strategies (including the optimal vaccine allocation strategy) we considered. We then describe the details of the optimization algorithm used to find the optimal vaccination strategy. Finally, we give details on the uncertainty analysis of our results.

### 2.1 Outbreak settings

Cholera has been present in Chad for decades [19]. In response to a large 2011 cholera epidemic in Chad, MSF worked closely with the Chadian Ministry of Health to try to limit the spread of the disease [20]. As part of that response, the MSF team and other health agencies collected case data in N’Djamena, the capital and largest city of Chad with a population of 993,500 in 2011, 19% of that are under five years old [20, 21]. It is unknown what triggered the epidemic, which resulted in 17,200 cases and 450 deaths, but it may be correlated with the occurrence of the rainy season [22, 23].

Between 2005 to 2012, surveillance had identified four cholera outbreaks in Maela, the largest refugee camp in Thailand with high population density and migration [7, 24]. The second largest of these outbreaks occurred in 2010 with 362 confirmed cases [25]. At this time, the camp was home to approximately 45,000 refugees, and 13% were children under five years old [7, 25].

Haiti comprises ten administrative departments, totaling 11 million inhabitants in 2010, with less than 12% under five years old [26]. In 2010 following a catastrophic earthquake, Haiti experienced its first recorded cholera outbreak in at least a century [27]. Poor access to water and sanitation in Haiti created a favorable condition for cholera to spread. Over the first two years of the epidemic, the Haitian Ministry of Public Health and Population reported more than 600,000 cases and over 7,000 deaths [28]. Since then, cholera has become endemic in Haiti [1].

### 2.2 The mathematical models of cholera transmission

Here we give a brief description of the models and highlight their differences. These models were used to analyze how different settings can alter the optimal use of OCV following an outbreak. For a complete description of the model structures and parameters, see the supplemental material (SM).

#### 2.2.1 Urban area: N’Djamena, Chad

We constructed a transmission model with vaccination and two age groups: those below five years old, and those who are at least five years old. In this model, individuals are either susceptible, exposed but not yet infectious, infectious with or without symptoms, or recovered—following an SEIR framework (Figure 1A). The model tracks the concentration of *V. cholerae* in the water reservoir. Transmission is seasonally forced and assumed to occur directly through contact with an infectious individual or indirectly through contact with contaminated water. We assume that asymptomatic infections are less infectious and shed less bacteria into the environment than symptomatic ones. Individuals may be unvaccinated or vaccinated with either one or two doses.

**Figure 1:**
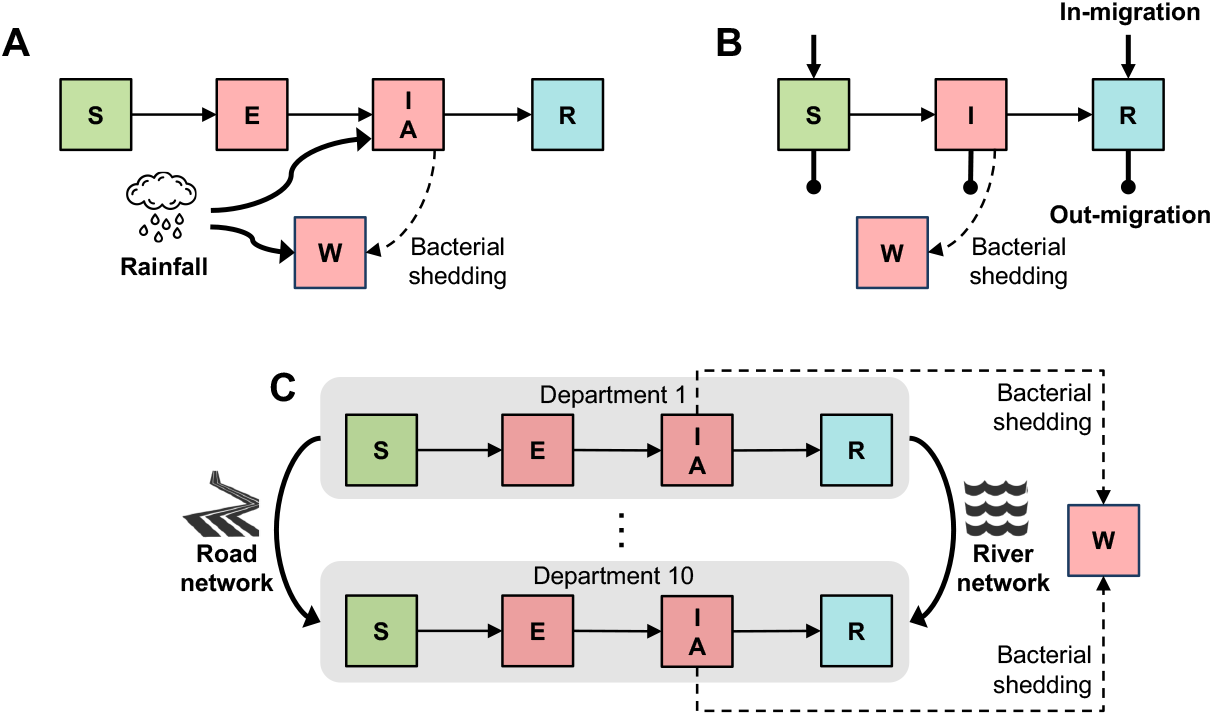
General framework of the models. A: N’Djamena, Chad, a densely populated city. B: Maela, a refugee camp. C: Haiti, where transmission is influenced by a road and water mobility network.

Using incidence data collected by MSF, we calibrated this model without age structure or vaccination to the 2011 epidemic in N’Djamena, Chad [29]. All parameters were fixed except for the direct transmission coefficient, environmental transmission coefficient, reporting ratio, and the number of recovered individuals at the start of the simulation. Details on the model calibration are found in the SM.

#### 2.2.2 Maela refugee camp, Thailand

We adapted a previously developed deterministic model of cholera transmission with two age groups that was calibrated to the 2010 epidemic in Maela to compare the impact of various vaccination campaigns [25]. In this model, individuals are either susceptible, infectious, or recovered (Figure 1B). They may be unvaccinated, once vaccinated, or twice vaccinated. We calibrated the two age groups (below five years old, and five years and older) to Maela demographics [7]. This model differs from the other two because individuals may enter and exit the population through births, in- and out-migration, and natural death.

#### 2.2.3 A region torn by natural disaster: Haiti

We used a previously developed meta-population model of cholera transmission in Haiti. Each node in the model represents one of the ten administrative departments in Haiti [26]. The departments are connected through a river network and a road network, and within each department, individuals transition through a SEIR framework (Figure 1C). Like in the model for Chad, infections may occur with or without symptoms. Environmental transmission is modulated by a sinusoidal function that represents the dry and rainy season. The mobility network (both rivers and roads) distinguishes this model from the models for Chad and Maela.

### 2.3 Vaccination campaign

For each of the three models, we built and implemented a vaccination campaign that considered two age groups, those under five years old and those at least five years old. We considered a “leaky” vaccine that reduces the probability of infection upon exposure (reducing susceptibility) [30]. The vaccine effectiveness varied by age and by the number of doses received (Table S1). Vaccine-induced immunity lasts an average of 4 years for two doses [31], and 2 years for one dose [32]. We assumed the duration of natural immunity was equal to the duration of vaccine-induced immunity after two doses.

We explored counterfactual vaccination scenarios following cholera epidemics under varying levels of vaccination coverage, with enough vaccine to cover 10 to 100% of the population with a single dose (5 to 50% of the population with two doses). For each setting, we considered vaccination strategies that may allocate zero, one or two doses to the different age groups. The vaccination strategies considered allocated

1. no vaccines at all (baseline);
2. two doses to both age groups proportional to population size (two-dose pro-rata);
3. one dose to both age groups proportional to population size (one-dose pro-rata);
4. two doses to those under five years old, and remainder allocated as one dose to those at least five years old (mixed);
5. one dose to those at least five years old (one-dose-over-five);
6. vaccine doses as found by our optimization routine (optimal).

We included the two-dose pro-rata strategy because this is the current standard, untargeted approach for mass OCV deployment worldwide [7, 8, 9, 10]. We considered two time horizons of one and three years. For each time horizon, we measured the performance of each vaccination strategy on three different metrics of disease burden: cumulative infections, cumulative symptomatic infections, and cumulative deaths.

Distribution of vaccine doses was assumed to take place over one week per round of vaccination, with 14 days between the first and second round [33]. We assumed that protection began immediately after the latest dose received, so that those who are allocated two doses were protected by the first dose until they received their second dose. Vaccination campaigns in Chad and Maela began two weeks after the start of simulation at 50 and 20 cumulative cases respectively, and in Haiti, at the start of simulation with 7300 cumulative cases, consistent with the initiation of surveillance incidence data [26].

### 2.4 Optimization

We combined our dynamic models of cholera transmission and vaccination for each setting with optimization algorithms to determine the optimal use of vaccine to minimize each disease metric. We used a previously developed optimization routine [34, 35] that works in two steps: first, it performs an exhaustive grid search that allows us to explore the entire variable space within a 5% margin of vaccine allocation, followed by a heuristic global algorithm that uses the results from the grid search as initial conditions (description of the optimization routine can be found in the SM).

### 2.5 Uncertainty analysis

Once a solution was deemed optimal, we performed uncertainty analysis by comparing this solution to the two-dose pro-rata, the one-dose pro-rata, mixed, and the one-dose-over-five strategies for 1000 parameter sets. We report here the mean and 95% uncertainty intervals (full details can be found in the SM).

## 3 Results

### 3.1 Optimal vaccine allocation by metrics of disease burden

In the next sections, we provide results considering a time horizon of one year. The optimal strategies to minimize cumulative infections, symptomatic infections, and deaths over one year were similar across Chad, Maela, and Haiti. When minimizing (total and symptomatic) infections, the optimal allocation strategies were largely identical in all three settings: Prioritize those at least five years old with a single dose of vaccine (Figure 2A, B, D, E, G, H). Once they have received one dose, the optimal strategies allocated any remaining vaccine (above 80% vaccination coverage; Figure 2) to those under five years old with two doses in Chad (A, B), as a second dose to those at least five years old in Haiti (G, H), or either of these depending on the metric in Maela (D, E). When minimizing deaths, the optimal strategies were equal to the ones to minimize symptomatic infections for Chad and Maela (Figure 2C, F). In contrast, in Haiti, the optimal strategy to minimize deaths prioritized the age groups in reverse order: Allocate two doses to children under five years old before distributing one dose to those at least five years old (Figure 2I).

**Figure 2:**
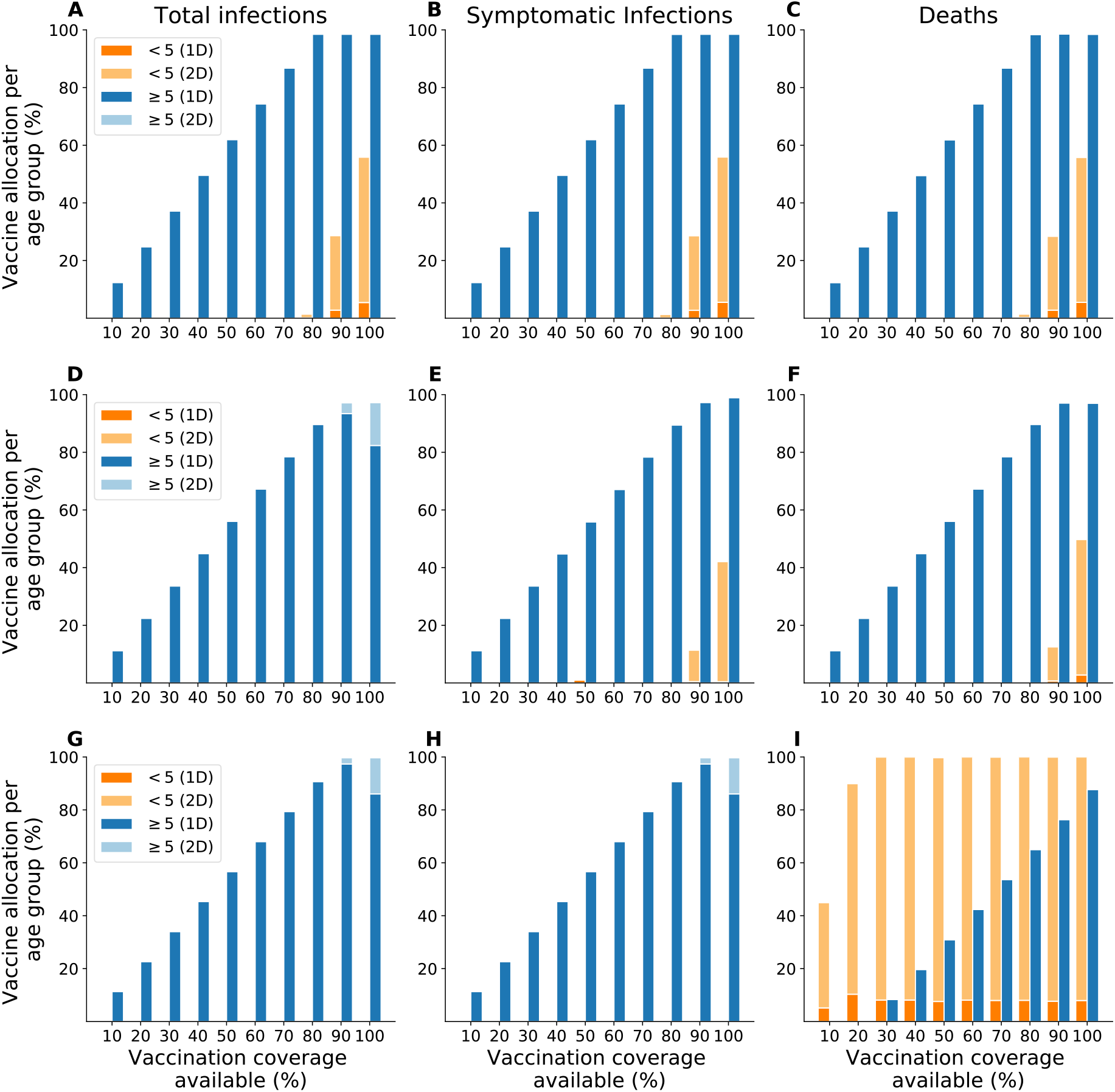
Optimal vaccine allocation strategies to minimize total infections, symptomatic infections, or deaths over one year. A–C: Chad. D–F: Maela. G–I: Haiti. We considered enough vaccine to cover 10–100% of the population with a single dose.

As expected, reactive vaccination in an outbreak setting can substantially curtail the epidemic. Without vaccination the prevalence of cases peaked at 23, 60, and 179 cases per 100,000 for Chad, Maela, and Haiti respectively (Figure 3). Using the optimal strategies that minimized symptomatic infections (Figure 2B, E, H), vaccination lowered the peak prevalence the most in Maela (Figure 3B) and least in Haiti (Figure 3C). At 50% coverage, these optimal strategies lowered the peak prevalence (per 100,000) to 10 in Chad, 12 in Maela, and 133 in Haiti (or by 50%, 80% and 26% respectively) (Figure 3). Next, we evaluated how the optimal strategies by objective performed against other vaccination strategies as measured by the percentage averted for the three metrics of disease burden: total infections, symptomatic infections, and deaths.

**Figure 3:**
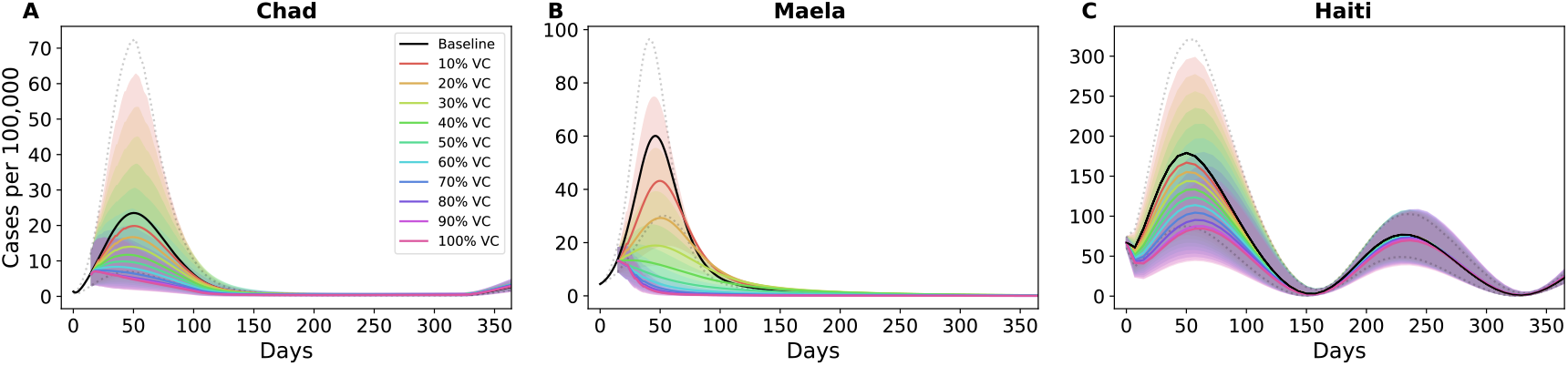
Prevalence of cases per 100,000 over one year. Prevalence of cases using an optimal vaccine allocation that minimized symptomatic infections over different vaccination coverage for A: Chad, B: Maela, and C: Haiti.

### 3.2 Performance of the optimal strategies in Chad, Maela, and Haiti

In Chad, the optimal strategies that minimized each metric of disease burden encompassed the one-dose-over-five strategy. These optimal strategies could have reduced up to 68% of total and symptomatic infections and 61% of deaths (Figure 4A–C). The optimal strategies minimizing total infections, symptomatic infections, and deaths maximally outperformed the two-dose pro-rata strategy at 80% coverage: by 23%, 23%, and 17% respectively. Compared to the one-dose pro-rata strategy, these optimal strategies averted up to 8% (80% coverage) more total and symptomatic infections and averted up to 6% (80% coverage) more deaths (Figure 4A–C).

**Figure 4:**
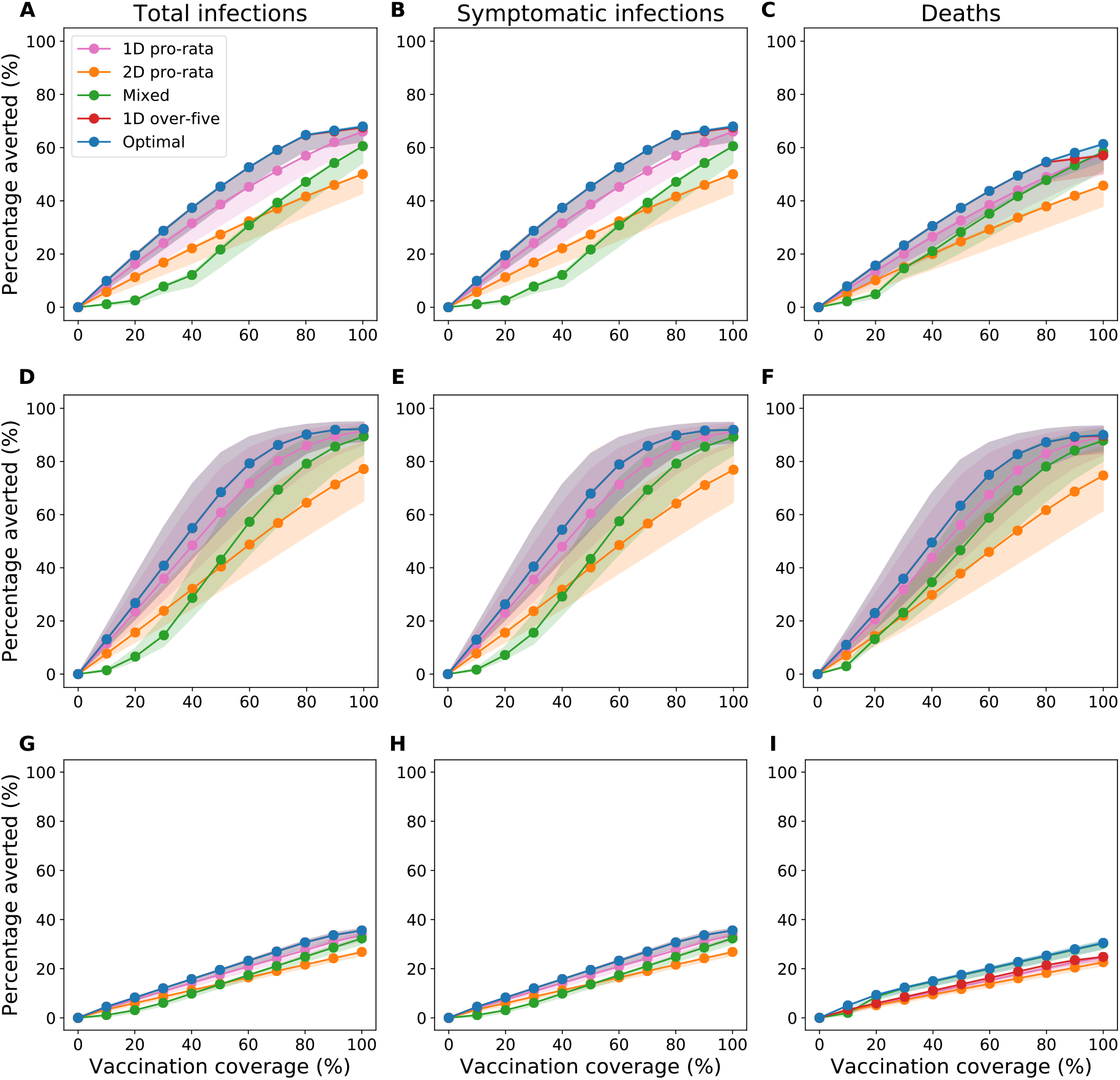
Reductions in the metrics of disease burden from the vaccination campaigns. Percentage of total infections (left), symptomatic infections (center), and deaths (right) averted over one year for A–C: Chad, D–F: Maela, and G–I: Haiti. We considered enough vaccine to cover 10–100% of the population with a single dose.

In Maela, the optimal strategies to minimize each metric also encompassed the one-dose-over- five strategy, outperforming all other vaccination strategies (Figure 4D–F). The optimal strategies for each objective had higher reductions than the two-dose pro-rata strategy by up to 30% (and higher than the one-dose pro-rata strategy by up to 8%). At 60% vaccination coverage, the optimal strategies for each objective had averted total infections, symptomatic infections, and deaths by at least 75%.

In Haiti, the optimal strategies reduced at most 36% of total and symptomatic infections and 31% of deaths (Figure 4G–I). The optimal strategies outperformed the two-dose pro-rata strategy by a small margin (up to 9% of total and symptomatic infections and 8% of deaths) and the one-dose pro-rata strategy by a smaller margin still (up to 3% of total and symptomatic infections and 6% deaths).

Among the three settings, Maela had the highest percentage of total infections, symptomatic infections, and deaths averted by the optimal strategies, followed by Chad and Haiti (Figure 4). For example, at 80% vaccination coverage in Maela, Chad, and Haiti, the optimal strategies that minimized deaths averted 87%, 55%, and 25% of deaths respectively (Figure 4C, F, I).

### 3.3 Comparison of the vaccination strategies across settings

The vaccination strategies that allocated a single dose of OCV outperformed the ones that allocated two doses for all three metrics and settings (Figure 4A–H), except for averted deaths in Haiti (I). The optimal strategies had the highest reductions for all metrics, followed by the one-dose pro-rata strategy (Figure 4A–H). The strategies allocating two doses (two-dose pro-rata strategy and mixed strategy) had smaller reductions: The two-dose pro-rata strategy outperformed the mixed strategy at lower vaccination coverage, while the latter outperformed the former at higher coverage. This crossover occurred at between 30% to 60% vaccination coverage (Figure 4).

Comparing the pro-rata vaccination strategies, one-dose outperformed a two-dose allocation, irrespective of setting, metric of disease burden, and vaccination coverage. For example, for averted deaths, the one-dose pro-rata strategy outperformed the two-dose pro-rata strategy by up to 22%, 12%, and 2% of deaths in Maela, Chad, and Haiti respectively (Figure 4C, F, I).

These reductions in disease burden show that early vaccination has the biggest impact. In Maela and Chad, where vaccination started early with respect to the epidemic curve, vaccination prevented up to 90% and 61% of deaths respectively, compared to 28% in Haiti, where vaccination began later.

### 3.4 Short-term versus long-term impact

We considered how the optimal strategies may change over a longer time horizon of 3 years. Recurring epidemics corresponded to seasonally forced transmission in both models for Chad and Haiti. In Maela, one case was re-seeded 1.5 years after the first identified case.

#### 3.4.1 Optimal strategies over a three year time horizon

When minimizing overall and symptomatic infections, the optimal strategies in Maela, Chad, and Haiti all prioritized individuals at least five years old (Figure 5A, B, D, E, G, H). In Maela and Chad, this age group was allocated a single dose when vaccine supply was low (below 40% vaccination coverage) and a mix of one and two doses with higher vaccine supply (Figure 5A, B, D, E). In Haiti, the optimal strategies allocated two doses to those at least five years old regardless of vaccine availability (Figure 5G, H).

**Figure 5:**
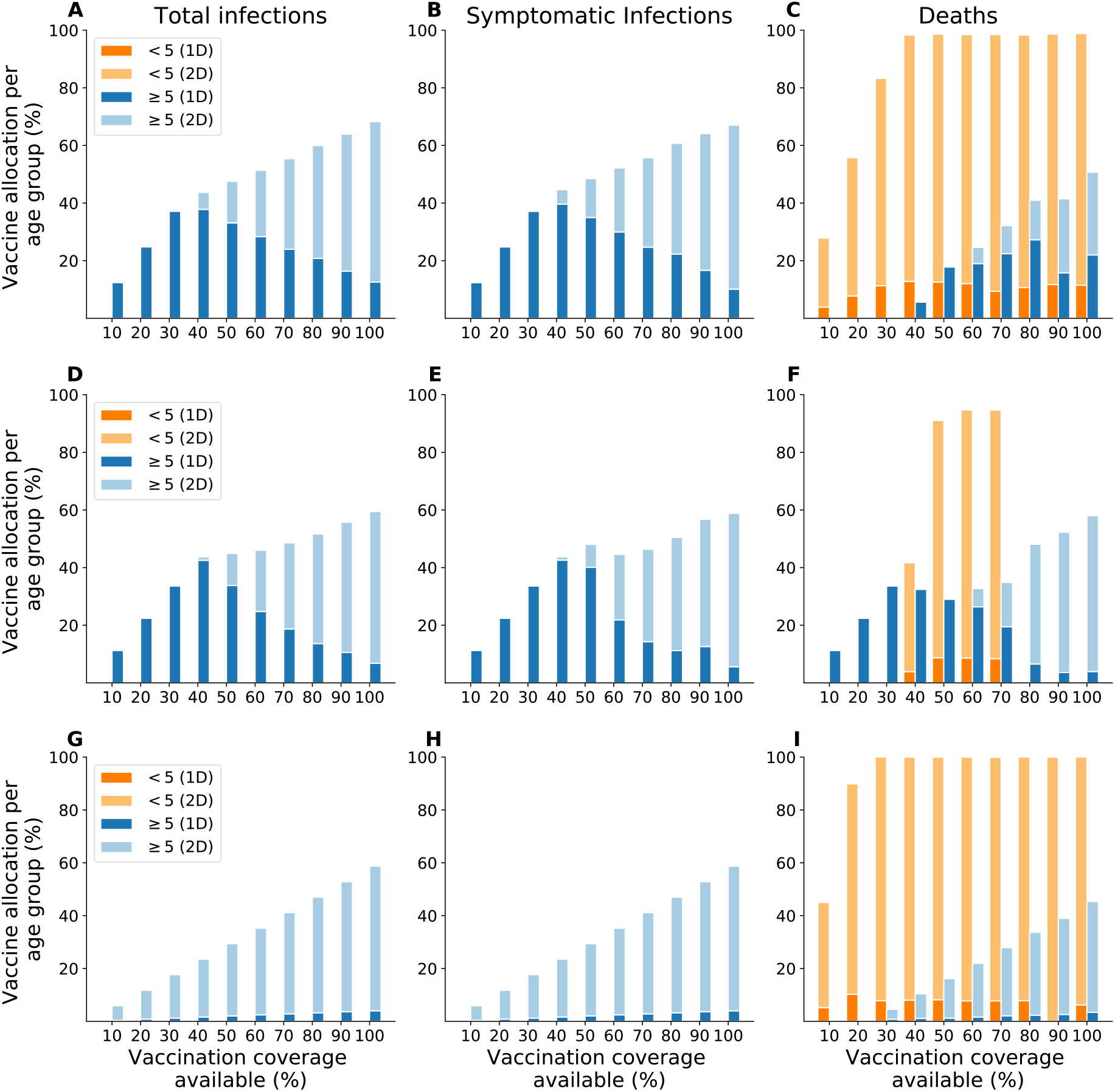
Optimal vaccine allocation strategies to minimize total infections (left), symptomatic infections (center), and deaths (right) over three years. A–C: Chad. D–F: Maela. G–I: Haiti. We considered enough vaccine to cover 10–100% of the population with a single dose.

When the objective was to minimize deaths over three years, the optimal strategies varied across Chad, Maela, and Haiti. In Chad, the optimal strategy was similar to the mixed strategy, prioritizing children under five years old with mostly two doses for all vaccination coverage. Once these children were vaccinated (above 30% vaccination coverage), the optimal strategy allocated the remaining vaccine to those at least five years old with a mix of one and two doses (Figure 5C).

In Haiti, the optimal strategy first targeted children under five years old with mostly two doses before allocating two doses to those at least five years old with any remaining vaccine (Figure 5I). In Maela, the optimal strategy prioritized the age group at least five years old with one dose (below 60% vaccination coverage) and with two doses (60% coverage and above), and allocated some vaccine to children under five years old (40% to 70% coverage) (Figure 5F).

#### 3.4.2 Performance of vaccination strategies over three years

The impact of the vaccination strategies was smaller at three years than at one year (Figure 6). In Maela, the re-introduced case did not produce any additional outbreaks, whereas there were multiple epidemic waves in Chad and Haiti. Across the settings, vaccination strategies over the longer term had the biggest impact in Maela, followed by Chad and Haiti (same order as over the shorter term).

**Figure 6:**
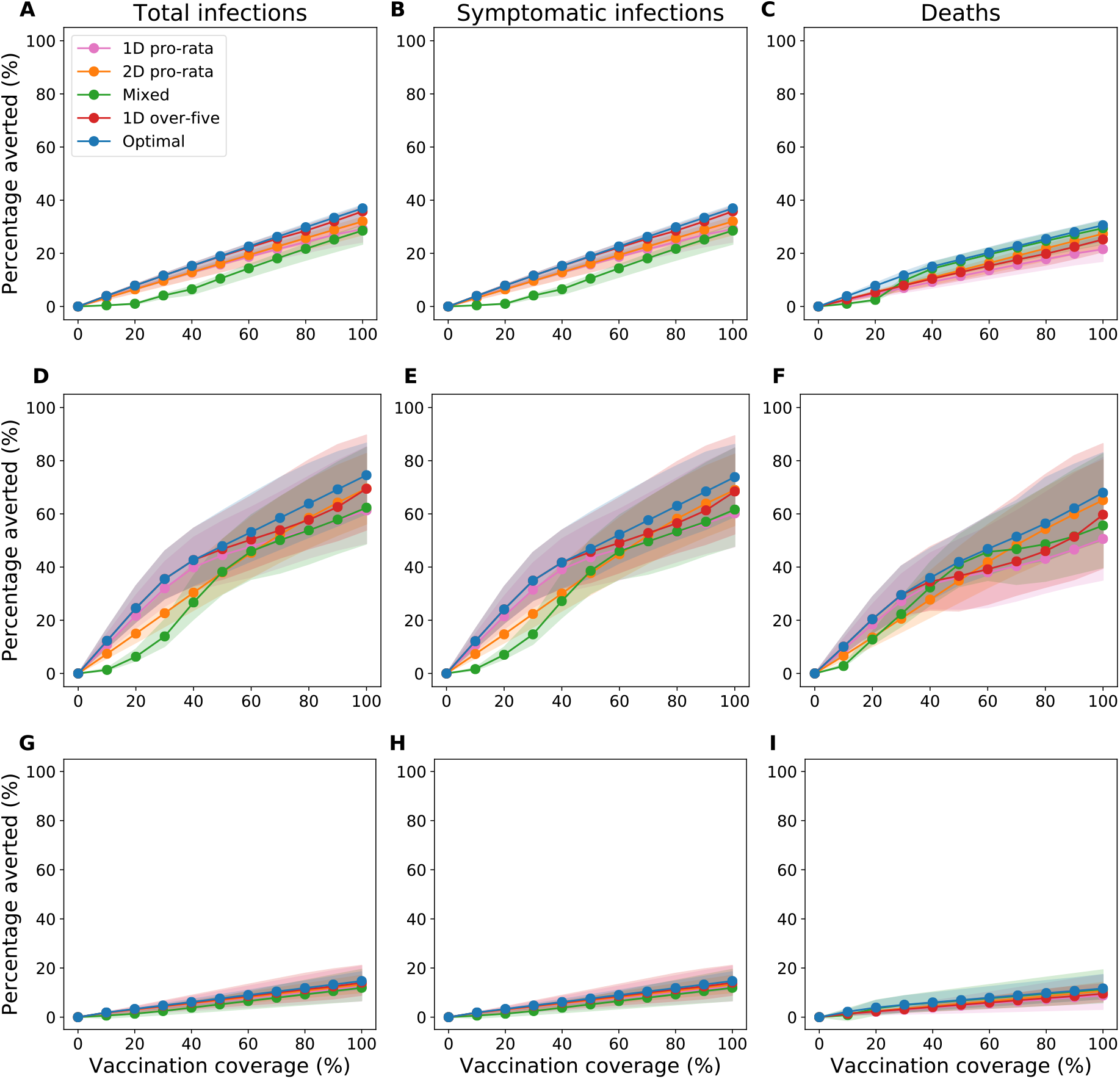
Reductions in metrics of disease burden from the vaccination campaigns over three years. Percentage of total infections (left), symptomatic infections (center), and deaths (right) averted over three years for A–C: Chad, D–F: Maela, and G–I: Haiti. We considered enough vaccine to cover 10–100% of the population with a single dose.

The performance differences over three years among various vaccination strategies were reduced from those over one year. When minimizing (total and symptomatic) infections over three years (Figure 6), the optimal strategies maximally outperformed the other strategies by 8% in Chad (A, B), 14% in Maela (D, E), and 2% in Haiti (G, H). When minimizing deaths, the optimal strategy at 80% vaccination coverage for Chad, Maela, and Haiti averted 25%, 56%, and 10% of deaths respectively (Figure 6C, F, I). At this vaccination coverage, the differences in deaths averted between the highest (optimal) and lowest (one-dose pro-rata) performing strategies were 8% in Chad, 13% in Maela, and 3% in Haiti. Over the longer term, the one-dose pro-rata strategy did not always outperform the two-dose pro-rata strategy. This is expected, as single-dose vaccination was assumed to wane after two years.

The optimal strategies over one and three years indicate that it is best to allocate one dose under limited vaccine supply in an outbreak setting for the highest reductions in disease and deaths. When more vaccine is available later, a second dose can be allocated.

### 3.5 A vaccine that reduces the likelihood of symptoms upon infection

In this section, we considered how the optimal strategies may change if a vaccine reduced the likelihood of having symptoms upon infection (disease-reducing) instead of one that reduced susceptibility to infection. We used the model calibrated for Chad.

#### 3.5.1 Optimal vaccination strategies

Over one year, the optimal strategies to minimize total infections, symptomatic infections, and deaths were the same for both vaccines (Figure S1), prioritizing a single dose to those at least five years old before allocating two doses to the younger age group. Over three years, the optimal strategies for each objective using a disease-reducing vaccine were slightly different to those using a susceptibility-reducing vaccine. When minimizing total and symptomatic infections for both vaccines, the optimal strategies for both vaccines prioritized a single dose to those at least five years old. For the disease-reducing vaccine, a second dose was allocated the same group at above 70% vaccination coverage (Figure S2A, B), whereas for the susceptibility-reducing vaccine, it was allocated sooner (at above 30% coverage) (Figure 5A, B). When minimizing deaths, the optimal strategies prioritized two doses to children under five years old for both vaccines (Figure S2C, Figure 2C). Any remaining vaccines were allocated to those at least five years old as a single dose (with the disease-reducing vaccine) or as a mix of one and two doses (susceptibility-reducing vaccine).

#### 3.5.2 Comparison of optimal strategies using two different vaccines

Here we compare the performance of the optimal strategies using the two different vaccines. When minimizing total infections, using a susceptibility-reducing vaccine outperformed using a disease-reducing vaccine by up to 23% and 19% over one and three years respectively (Figure 4A, S3A, 6A, S4A). When minimizing symptomatic infections and deaths, however, the optimal strategies using a disease-reducing vaccine outperformed ones using a susceptibility-reducing vaccine by up to 2% over one year and 12% over three years (Figure 4B–C, S3B–C, 6B–C, S4B–C).

#### 3.5.3 Comparison of optimal strategies to other vaccination strategies

Here we compare the optimal strategies using the disease-reducing vaccine with other vaccination strategies. There was no qualitative difference in the performance ranking among the various vaccination strategies over one year between the two vaccines. The one-dose strategies still broadly outperformed two-dose strategies for all metrics of disease burden over one year and for averting total and symptomatic infections over three years (Figure S3, S4). The differences in performance between the strategies with the highest and lowest reductions for total infections, symptomatic infections, and deaths were up to 17%, 24%, and 17% over one year respectively (or 4%, 11%, and 10% over three years respectively) (Figure S3, S4).

## 4 Discussion

We paired mathematical models of cholera transmission and vaccination with optimization algorithms to determine the optimal vaccination strategies and their impact on various metrics of disease burden. We explored vaccination campaigns that included the use of one dose of OCV, which is half of the recommended two-dose regimen. We considered two target populations by age (under five years old, and five years and older), different levels of vaccination coverage, three distinct cholera outbreak settings, and a time horizon of one and three years. We compared these optimal strategies to other vaccination strategies.

We found that strategies allocating one dose of vaccine to those five years and older reduced the most cumulative infections, symptomatic infections, and deaths for all three outbreak settings in Chad, Maela, and Haiti over the short term. In Haiti, the optimal allocation strategies also fully protected children under five years old with two doses to minimize deaths. Our results suggest that over a shorter term (one year), strategies with a one-dose allocation to those aged five years and older, for whom the one-dose vaccine is more effective, outperform ones allocating two doses irrespective of setting, metric of disease burden, and vaccination coverage. Further, our results show that if reactive vaccination campaigns start when there are already a significant number of cases (Haiti in our settings) then it is still optimal to give one dose to those older than five, but to also give full protection to the youngest children. This would stretch the vaccine supply considerably, and of course, once vaccine supply is replenished, vaccination campaigns with a second dose for long-term protection are desired.

A modeling study has found that a one-dose vaccination campaign would avert 1.1 to 1.2 times as many cases and deaths as a two-dose campaign in an outbreak setting over the short term [36].

This is consistent with our finding that optimal strategies for each objective, which allocated a mix of one and two doses, would avert up to 1.2 to 1.8 times as many cases and deaths as a two-dose pro-rata strategy over the course of one year. Similarly, the one-dose pro-rata strategy would avert to 1.5 times as many cases and deaths as the two-dose pro-rata strategy across the three outbreak settings. Different to their study, we used two age groups and age-specific case fatality rates. In particular, we assumed that the case fatality rates for children under five years old were about 5 times higher than individuals who were older [37]. Another modeling study analyzed the impact of vaccination campaign timing and one-dose vaccine effectiveness and found that timing had a bigger impact on case counts than one-dose effectiveness [25]. This is consistent with our finding that the impact of vaccination was smallest in Haiti, where vaccination began later during the exponential phase of the epidemic.

The similar optimal vaccination strategies across three different settings provide reassurance that these results are robust to a range of transmission dynamics. However, there are limitations to our work. As with all mathematical models, our models have simplifying assumptions on the true transmission dynamics and use parameter values that are subject to some uncertainty. In particular, we assumed homogeneity in mixing patterns and probability of a symptomatic case given infection. We calibrated the Chad model to incidence data. However, the burden of cholera is under-reported [38, 39]. Countries may suppress reports of cholera cases due to the perceived influence cholera has on tourism and export industries [40]. Additionally, the WHO case definition for cholera currently excludes children under five years old [41]. This can further obscure the true burden of cholera, especially in areas where children face increased cholera incidence. Importantly, the case fatality ratio for cholera differs by age and geography. We assumed a fixed multiplier that increased childhood mortality. In reality, this multiplier is likely to change for different regions with different healthcare systems and access to clean water. There is some uncertainty around the vaccine efficacy of one dose of OCV. Trials of one-dose vaccine efficacy of OCV have demonstrated good protection for individuals over five years old, and low to no protection for children below five years old in Bangladesh where cholera is endemic [4, 5]. Understanding how that protection translates to other settings, such as in Haiti where cholera is newly endemic, should be a priority.

Despite the counterfactual nature of our vaccination campaigns, our results can provide a basis for future cholera outbreaks, especially under settings with poor water infrastructure and constrained resources. We explored hypothetical vaccination campaigns that took place two weeks after the first case was identified (Chad and Maela) and during the exponential phase of the epidemic (Haiti). In reality, in emergency situations, the time from vaccine request to vaccine delivery are often long [33]. In emergency situations, the median time between the first laboratory confirmation (or occurrence of a humanitarian emergency) to one week after the first round of vaccination when immunity develops was 66 days [33]. Our results point to the need to reduce these lag times in order to increase the impact of vaccination campaigns. A one-dose vaccine allocation would reduce this logistical hurdle, and it would not preclude a second dose once there is more vaccine supply.

## Data Availability

All data produced in the present study are available upon reasonable request to the authors.

## Acknowledgements

We thank Joshua Havumaki, Rafael Reza, Marisa Eisenberg, and Andrew Azman for providing access to research-related materials that were key to our analysis.

## Supplemental Figures

**Figure S1:**
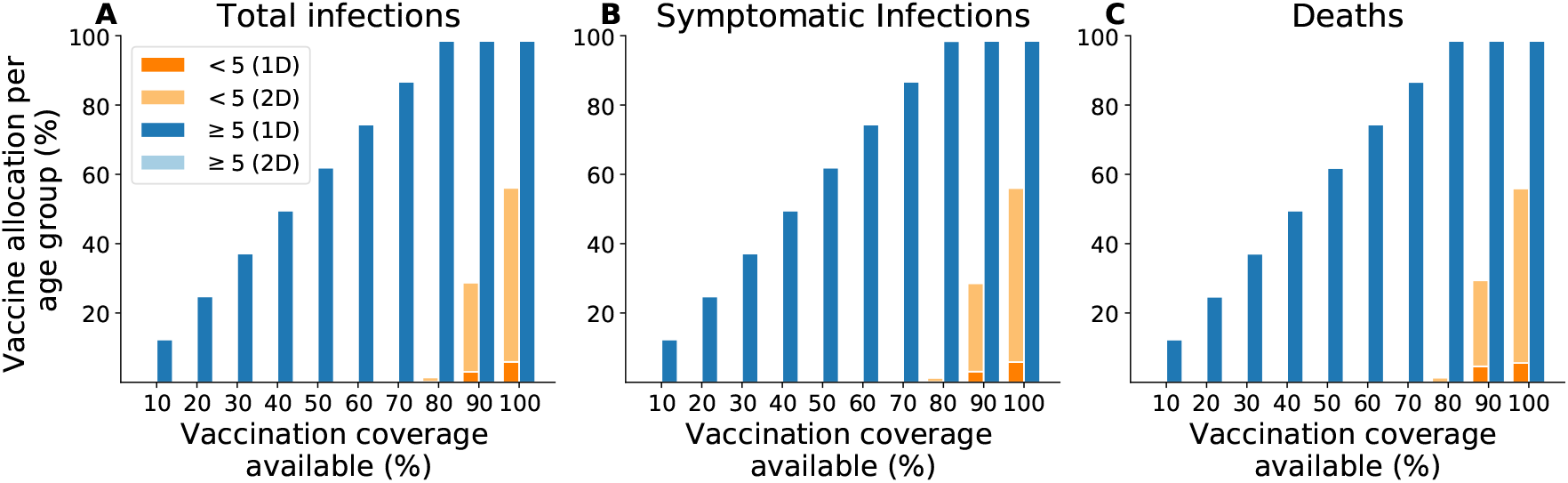
Optimal vaccine allocation strategies for different goals in Chad over one year using a vaccine that reduces the probability of symptoms upon infection. A: total infections. B: symptomatic infections. C: deaths. We considered enough vaccine to cover 10–100% of the population with a single dose.

**Figure S2:**
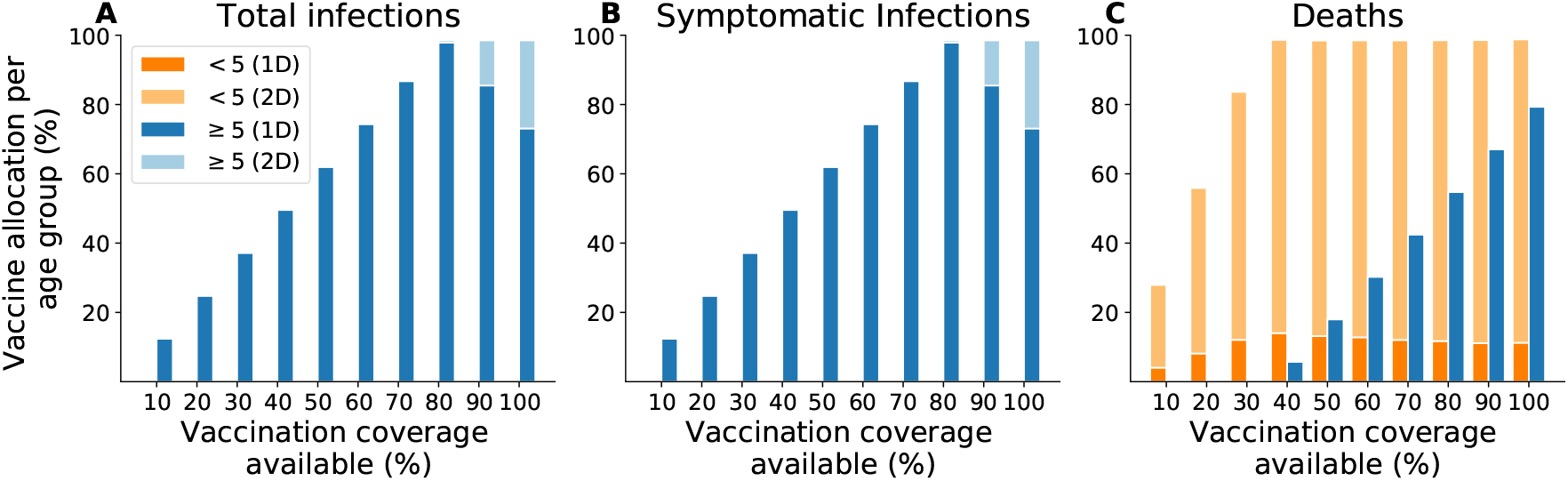
Optimal vaccine allocation strategies for different goals in Chad over three years using a vaccine that reduces the probability of symptoms upon infection. A: total infections. B: symptomatic infections. C: deaths. We considered enough vaccine to cover 10–100% of the population with a single dose.

**Figure S3:**
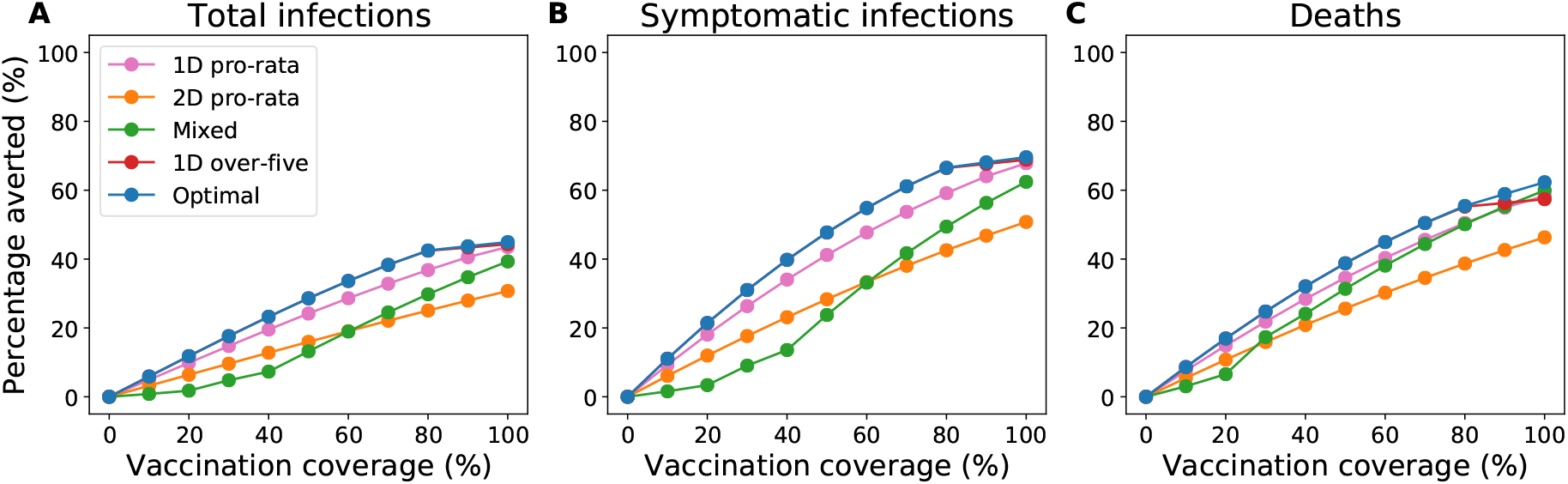
Reductions in metrics of disease burden from vaccination campaigns over one year with a vaccine that reduces the probability of symptoms upon infection. We considered enough vaccine to cover 10–100% of the population with a single dose.

**Figure S4:**
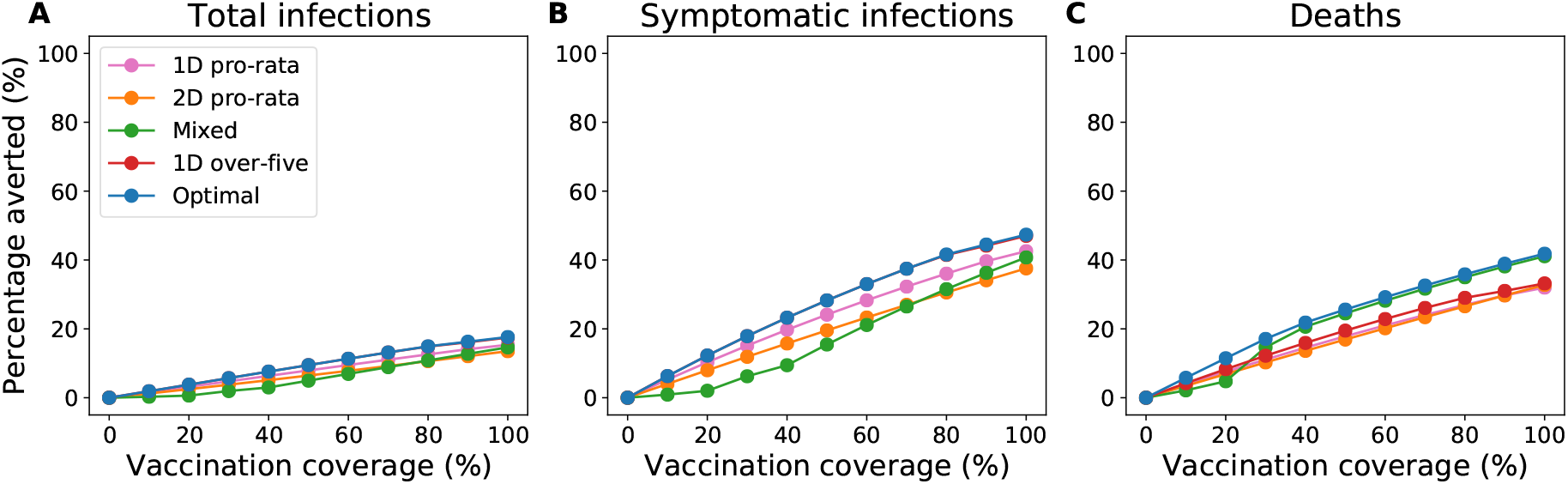
Reductions in metrics of disease burden from vaccination campaigns over three years with a vaccine that reduces the probability of symptoms upon infection. We considered enough vaccine to cover 10–100% of the population with a single dose.

## Appendix The models

### The urban city model: N’Djamena, Chad

We constructed a transmission model to simulate a cholera outbreak in an urban city and calibrated it to the 2011 cholera outbreak in N’Djamena, Chad (Figure S5). Susceptible individuals (*S*) are infected through contact with an infectious individual or with a water source. Upon infection, the individual becomes exposed (*E*) where they are infected but not yet infectious for an average 1*/γ*_*E*_ days. A fraction *k* of these exposed individuals will experience symptoms and transition to the infectious with symptoms (*I*) class. The remaining fraction (1 − *k*) will be asymptomatically infectious (*A*). Both infectious classes shed bacteria into the environment (water source). We assume that asymptomatic infectious individuals have a reduced degree of infectiousness *b*_*A*_ and reduced degree of bacterial shedding *b*_*µ*_ relative to symptomatic individuals. Natural immunity is assumed to last 1*/σ* (Erlang distributed) until the recovered individual becomes fully susceptible to infection again.

Environmental or waterborne transmission is measured by the amount of bacteria shed into the water reservoir (*W*). An infectious individual sheds bacteria in the water at rate *µ*, and bacteria decays at a rate *δ*. The force of infection is the sum of direct transmission by contact with an infectious individual and indirect transmission by contact with contaminated water, as measured by *β* and *β*_*W*_ respectively. Transmission by water is modulated by rainfall, which has been shown to be an important factor in cholera epidemics [18, 17]. A description of the model parameters is found in Table S2.

### The refugee camp model: Maela, Thailand

We extended a previously developed transmission model of cholera in Maela, the largest refugee camp in Thailand [25] (Figure S6). The formulation of the model is explained in detail in the work of Havumaki et al. [25]. Whereas the original version of the model used an exponentially distributed duration of immunity, immunity waned over two recovered compartments leading to an Erlang distributed duration of immunity. A table of the model parameters is found in Table S3.

### The region torn by natural disaster model: Haiti

We used a previously developed meta-population model of cholera transmission in Haiti that consists of ten administrative departments [26] (Figure S7). Details of the model are found in Lee et al. [26]. We made no changes to the transmission dynamics of this model. Model parameters are found in Table S4.

**Figure S5:**
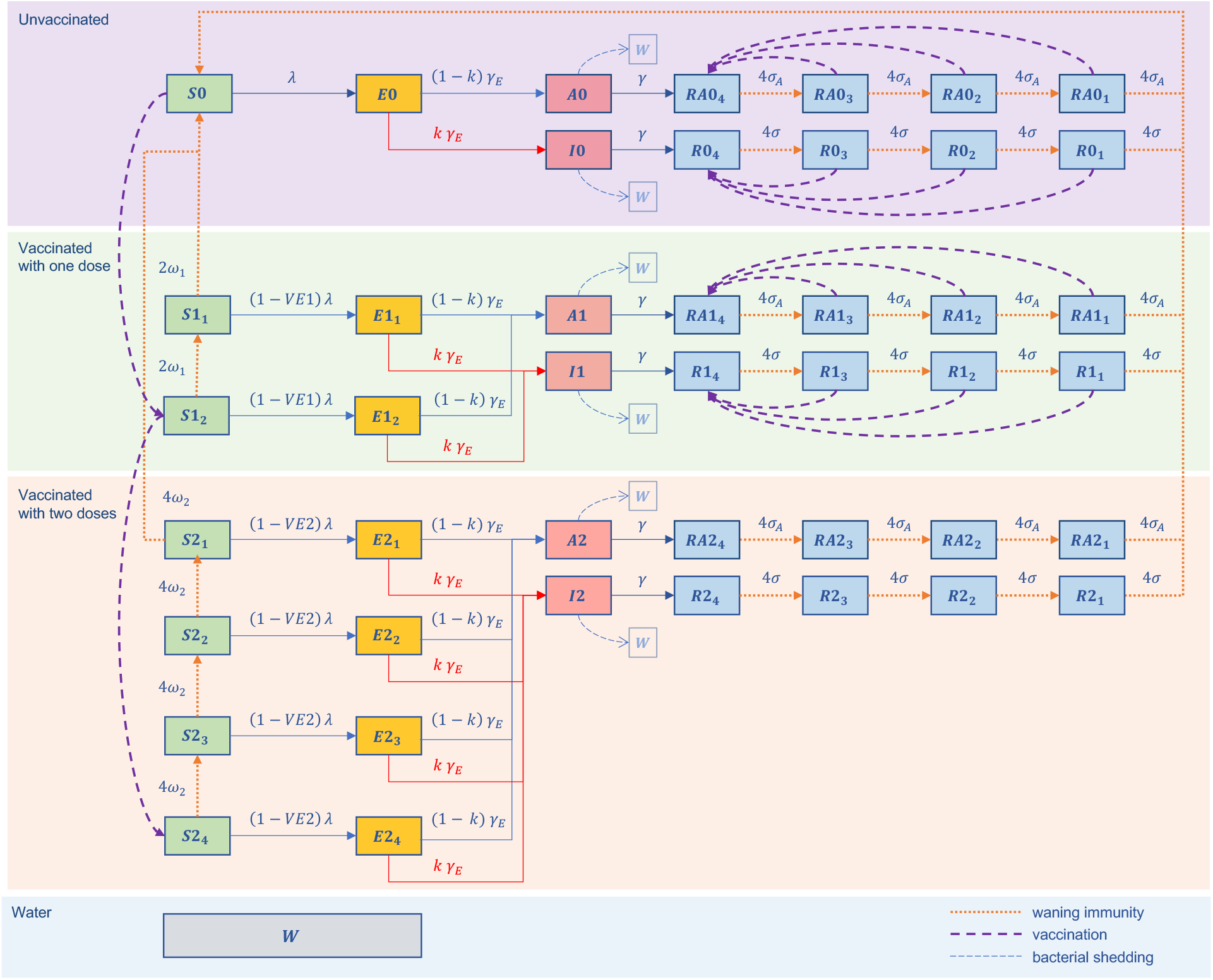
Diagram of the model of cholera transmission and vaccination in Chad.

**Figure S6:**
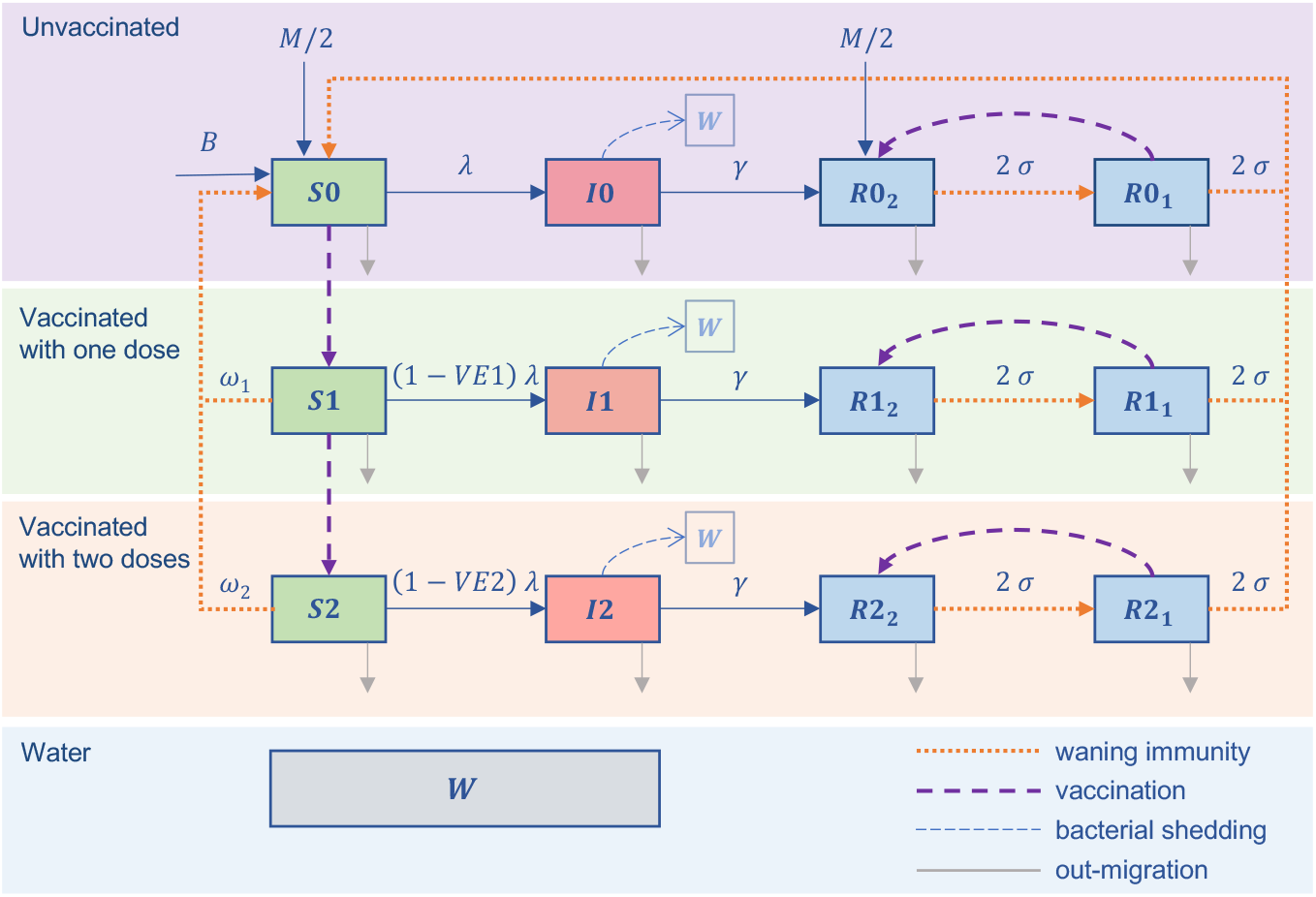
Diagram of the model of cholera transmission and vaccination in Maela.

**Figure S7:**
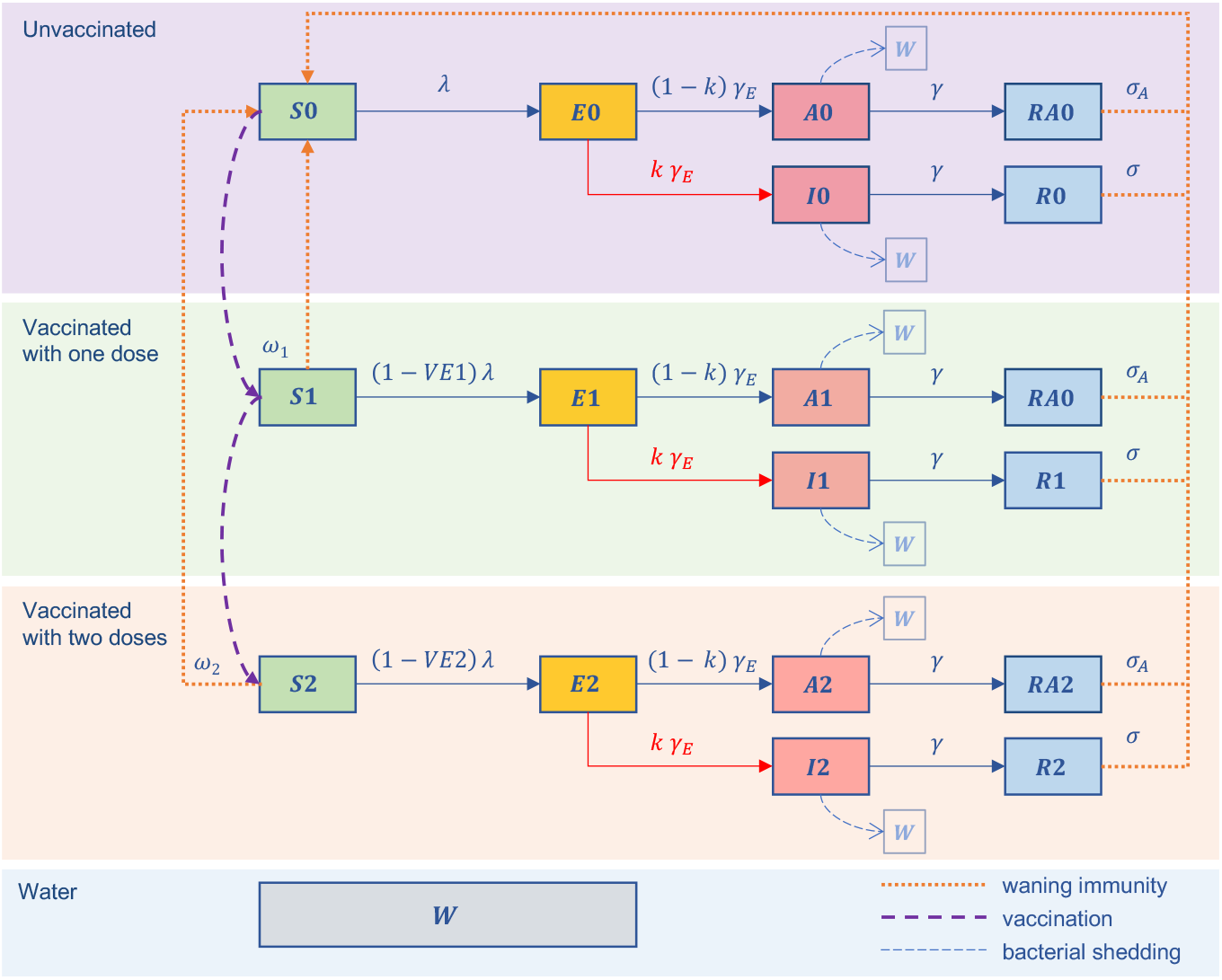
Diagram of the model of cholera transmission and vaccination in Haiti.

### Model Equations

The system of equations for the Chad model (age group omitted for clarity) are:

Unvaccinated:

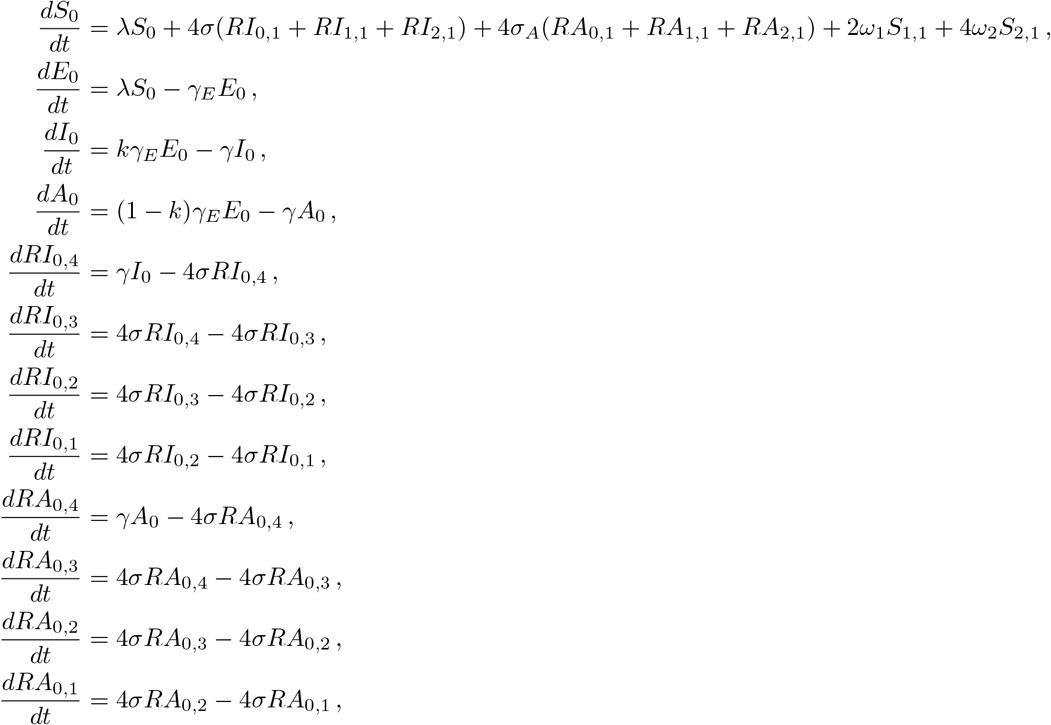

Vaccinated with one dose:

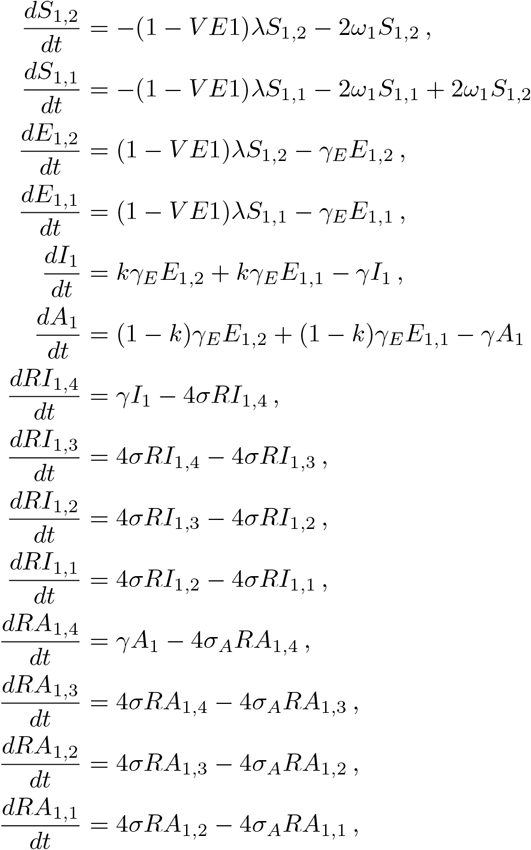

Vaccinated with two doses:

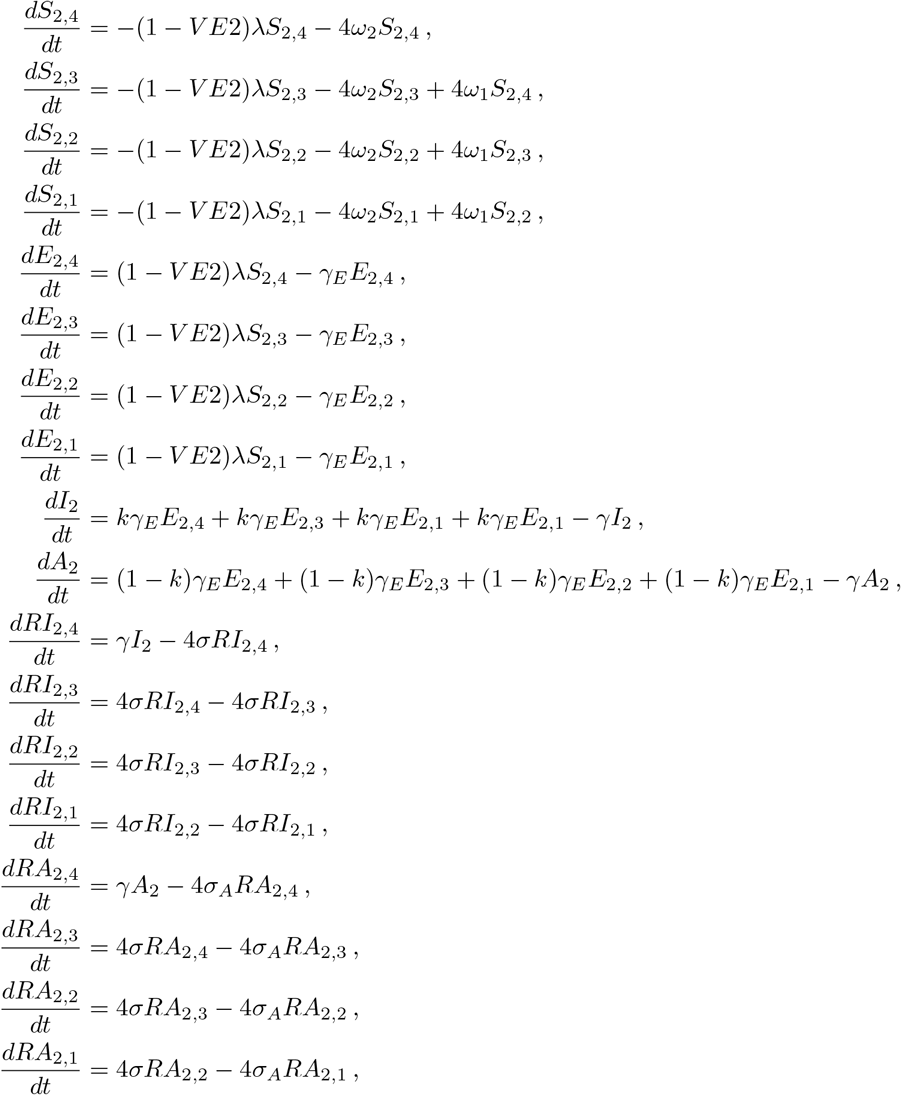

Water:

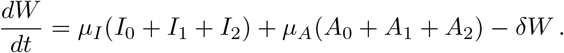

The force of infection *λ* is given by

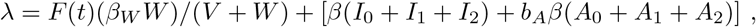

where

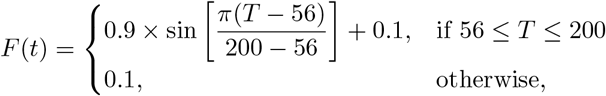

**Figure S8:**
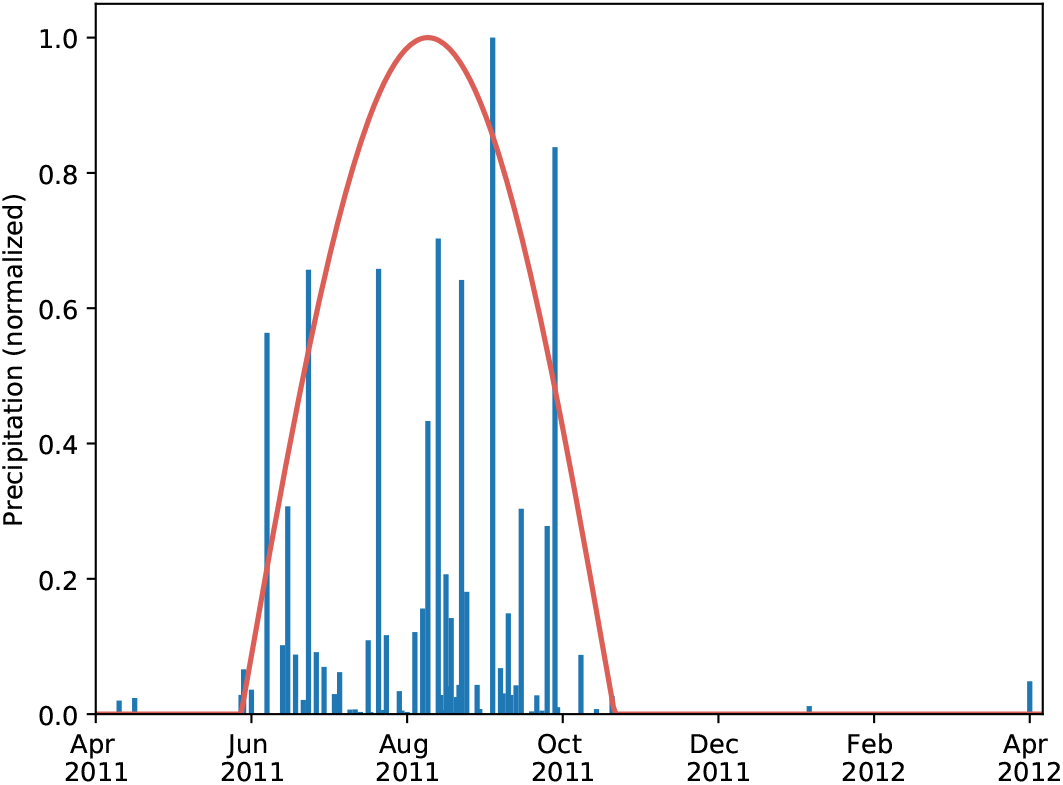
Representation of the rainfall in Chad. Environmental transmission is modulated by a constant during the dry season (December to June) and by a sinusoidal function during the rainy season (June to November).

*T* = (*t* − 365*n*) and the discrete year *n* = (0, 1, 2, …). The model diagram is found in Figure S5.

We obtained daily precipitation data for N’Djamena from April 2011 to April 2012 from the Tropical Rainfall Measuring Mission Multi-satellite Precipitation Analysis (Version 7) [42]^1^. During the rainy season between June and October, we modeled rainfall with a sine function (Figure S8).

### Model calibration: Chad

Daily incidence data for the 2011 cholera outbreak in N’Djamena, Chad were collected by Médecins Sans Fronti`eres, and its details are found in [29]. The data spanned 232 days from July to November 2011. We fitted the model of Chad without age structure or vaccination using particle swarm optimization (pyswarms package in Python) [43, 44]). All model parameters were fixed except for four: person-to-person transmission, environment-to-person transmission, reporting ratio, and the number of recovered individuals at the start of simulation. In order to focus on the exponential phase of the outbreak, we fit the model from Day 100 of the data when the new cases increased exponentially. We assumed the cases from the first 100 days of the epidemic (1551 cumulative cases) had recovered and used that as a lower bound for number of recovered individuals at the start of simulation. The calibrated model reproduced the epidemic curve of the 2011 epidemic in Chad (Figure S9). All parameter values, including the fitted ones, are shown in Table S2 for Chad.

**Figure S9:**
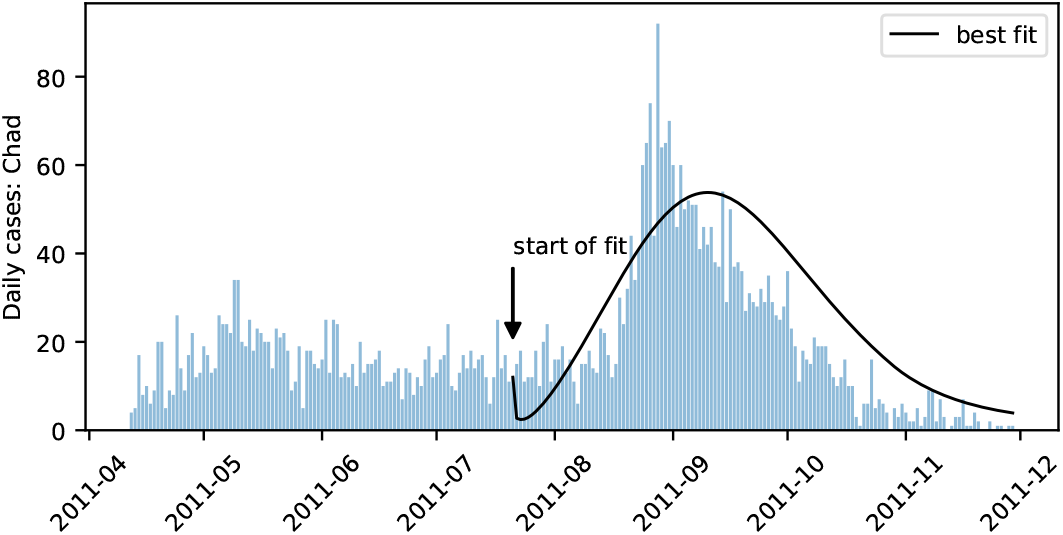
Incidence data from Chad and the model fit without vaccination.

### Vaccination dynamics

For each objective, the optimal allocation strategy determines how many doses to allocate to the two age groups that comprise the population (children under 5 years old, and those at least 5 years old). In the meta-population model of Haiti comprising ten administrative departments, we assume that the number of doses allocated per department is proportional to the population size of that department.

For a given optimal vaccine allocation strategy, the vaccination campaign is implemented in the following way. First, every individual allocated at least one dose receives a dose of vaccine. Then, those who are allocated two doses are vaccinated with their second dose. If the proposed number of vaccines allocated to an age group exceeded the population size of that group, the extra vaccines are assumed to be wasted.

In the vaccination campaign, available vaccine doses are distributed among susceptible and recovered individuals in proportion to their population size (pro-rata). Where the duration of immunity was Erlang-distributed (in Chad and Maela), recovered individuals who receive a vaccine have their immunity boosted. Where duration of immunity was exponentially distributed (in Haiti), only susceptible individuals were vaccinated. After an individual recovers from infection, immunity may wane.

### Uncertainty Analysis

We assessed the uncertainty in the output measures of disease burden (percentage of cumulative infections averted, percentage of cumulative symptomatic infections averted, and percentage of cumulative deaths averted) by performing 1,000 simulations each with different parameter sets. Because each model had different features, the parameters that varied for this part of the analysis also differed. For Chad and Haiti, we sampled pre-determined gamma distributions with means given in Table S2 and S4 for the mean latent period and the mean infectious period. For the direct transmission coefficient, environmental transmission coefficient, relative infectiousness of asymptomatic individuals, relative shedding of asymptomatic individuals, and fraction of infections that are symptomatic, we used truncated normal distributions with means and ranges given in Table S2 and S4. For Maela, we sampled the infectious period from gamma distributions with means provided in Table S3; the direct and environmental transmission coefficients and the reporting ratio for those at least five years old from truncated normal distributions with means and ranges specified in Table S3. The reporting ratio for those under five years old was calculated with a multiplier (1.37 times higher). The shaded areas presented in the figures were obtained by removing the top and bottom 2.5% of the simulations.

### Optimization

We developed a heuristic optimization procedure consisting of two stages allowing us to explore the entire vaccine space [34, 35]). We give a brief description here. The decision variable for the optimization problem is the proportion of the total vaccine available allocated to each of four groups (two age groups receiving one or two doses):

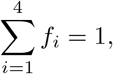

where *f*_*i*_ ≥ 0 is the fraction of the total available vaccine allocated to group *i*. The optimization procedure is as follows. In the first stage, we computed a mesh on the feasible set in 5% increments. We evaluated the metric (objective function) at each point on this mesh, to find the *N* best points (that is, those points with the *N* smallest metric values). In the second stage, we then randomly sampled *N* additional points from the Dirichlet distribution [50]. We use the 2*N* points as initial points for an unconstrained optimization method (Nelder-Mead [51, 52] or Particle Swarms [43, 44]), plus four additional starting points: one-dose-pro-rata, two-dose-pro-rata, one-dose-over-five, and the mixed strategy. Within the function evaluation step of the unconstrained method, the decision variable is corrected to satisfy *f*_*i*_ ≥ 0 for *i* = 1, …, 4 and 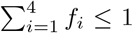. The “optimal solution” is declared to be the best of these solutions.

**Table S1:** Description of parameters related to the vaccine and disease metrics.

| Parameter | Meaning | Value < 5 | Value $\geq 5$ | Source |
| --- | --- | --- | --- | --- |
| **Vaccine efficacy:** |  |  |  |  |
| $VE1$ | reduced susceptibility to infection after 1 dose | 0.08 | 0.575 | [4, 5] |
| $VE2$ | reduced susceptibility to infection after 2 doses | 0.42 | 0.72 | [3] |
| $1/\omega_1$ | mean duration of immunity from 1 dose | 2 years | 2 years | [32] |
| $1/\omega_2$ | mean duration of immunity from 2 doses | 4 years | 4 years | [31, 32, 3] |
| **Case fatality rate:** |  |  |  |  |
|  | Chad: | 0.21014 | 0.038 | [37, 1] |
|  | Thailand: | 0.04977 | 0.009 | [37, 45] |
|  | Haiti: | 0.07742 | 0.014 | [28] |

**Table S2:** Description of parameters used in the model of Chad, unless otherwise specified. (p = person; t = days)

| Parameter | Meaning | Value (Range) [Units] | Source |
| --- | --- | --- | --- |
| **Chad:** |  |  |  |
| $\beta$ | direct transmission coefficient | $3.31 \times 10^{-6}$ ($2.48 \times 10^{-6}$, $4.13 \times 10^{-6}$) [$\frac{1}{p \cdot t}$] | fitted |
| $\beta_W$ | environmental transmission coefficient | $1.43 \times 10^{-3}$ ($1.07 \times 10^{-3}$, $1.79 \times 10^{-3}$) [$\frac{1}{p \cdot t}$] | fitted |
| $1/\delta$ | bacterial decay rate | 21 [t] | [46] |
| $1/\gamma_E$ | mean latent period | 2 [t] | [47] |
| $\mu_I$ | bacterial shedding rate of symptomatic individuals | 575 [$\frac{\text{cells/mL}}{p \cdot t}$] | |
| $\mu_A$ | bacterial shedding rate of asymptomatic individuals | $0.01 \times 575$ [$\frac{\text{cells/mL}}{p \cdot t}$] | [47] |
| $b_A$ | reduced infectiousness of asymptomatic infections | 0.01 (0.0075, 0.0125) | [47] |
| $b_\mu$ | reduced bacterial shedding of asymptomatic infections | 0.01 (0.0075, 0.0125) | [47] |
| $1/\sigma$ | mean duration of natural immunity | $4 \times 365$ [t] | — |
| $r$ | reporting ratio | 0.08987 | fitted |
| $k$ | proportion of infections that are symptomatic | 0.1 (0.05, 0.15) | [48, 49] |
| $1/\gamma$ | mean infectious period | 4 (2, 6) [t] | [29] |
| $V$ | bacteria concentration yielding 50% of catching infection | 10,000 [cells/mL] | [46] |
| $R(0)$ | recovered individuals at the start of simulation | 198,015 [p] | fitted |
| $p_{\text{under}5}$ | proportion of population under 5 years old | 0.193 | [21] |

**Table S3:**
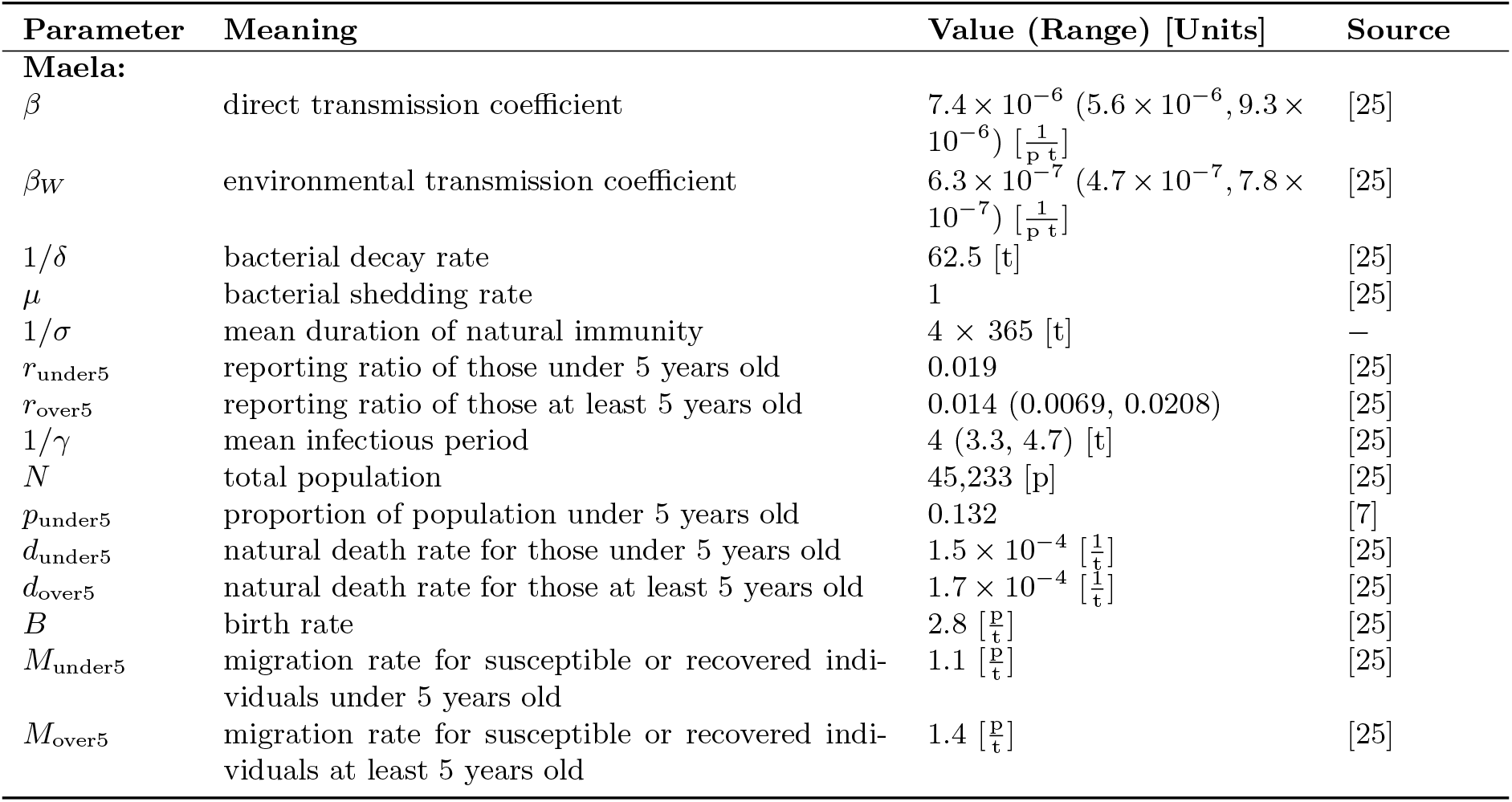
Description of parameters used in the model of Maela, including the values used, unless otherwise specified. (p = person; t = days)

| Parameter | Meaning | Value (Range) [Units] | Source |
| --- | --- | --- | --- |
| **Maela:** |  |  |  |
| $\beta$ | direct transmission coefficient | $7.4 \times 10^{-6}$ ($5.6 \times 10^{-6}, 9.3 \times 10^{-6}$) [$\frac{1}{p \cdot t}$] | [25] |
| $\beta_W$ | environmental transmission coefficient | $6.3 \times 10^{-7}$ ($4.7 \times 10^{-7}, 7.8 \times 10^{-7}$) [$\frac{1}{p \cdot t}$] | [25] |
| $1/\delta$ | bacterial decay rate | 62.5 [t] | [25] |
| $\mu$ | bacterial shedding rate | 1 | [25] |
| $1/\sigma$ | mean duration of natural immunity | $4 \times 365$ [t] | — |
| $r_{\text{under5}}$ | reporting ratio of those under 5 years old | 0.019 | [25] |
| $r_{\text{over5}}$ | reporting ratio of those at least 5 years old | 0.014 (0.0069, 0.0208) | [25] |
| $1/\gamma$ | mean infectious period | 4 (3.3, 4.7) [t] | [25] |
| $N$ | total population | 45,233 [p] | [25] |
| $p_{\text{under5}}$ | proportion of population under 5 years old | 0.132 | [7] |
| $d_{\text{under5}}$ | natural death rate for those under 5 years old | $1.5 \times 10^{-4}$ [$\frac{1}{t}$] | [25] |
| $d_{\text{over5}}$ | natural death rate for those at least 5 years old | $1.7 \times 10^{-4}$ [$\frac{1}{t}$] | [25] |
| $B$ | birth rate | 2.8 [$\frac{p}{t}$] | [25] |
| $M_{\text{under5}}$ | migration rate for susceptible or recovered individuals under 5 years old | 1.1 [$\frac{p}{t}$] | [25] |
| $M_{\text{over5}}$ | migration rate for susceptible or recovered individuals at least 5 years old | 1.4 [$\frac{p}{t}$] | [25] |

**Table S4:** Description of parameters used in the model of Haiti, unless otherwise specified. (p = person; t = weeks)

| Parameter | Meaning | Value (Range) [Units] | Source |
| --- | --- | --- | --- |
| **Haiti:** |  |  |  |
| $\beta$ | direct transmission coefficient | $9.9 \times 10^{-7}$ ($7.45 \times 10^{-7}, 1.24 \times 10^{-6}$) [$\frac{1}{p \cdot t}$] | [26] |
| $\beta_W$ | environmental transmission coefficient | 0.04 (0.03, 0.05) [$\frac{1}{p \cdot t}$] | [26] |
| $1/\delta$ | bacterial decay rate | 3 [t] | [26] |
| $1/\gamma_E$ | mean latent period | 0.18 (0.014, 0.550) [t] | [26] |
| $\mu_I$ | bacterial shedding rate of symptomatic individuals | 3982.08 [$\frac{\text{cells/mL}}{p \cdot t}$] | [26] |
| $\mu_A$ | bacterial shedding rate of asymptomatic individuals | $10^{-7} \times \mu$ [$\frac{\text{cells/mL}}{p \cdot t}$] | [26] |
| $b_A$ | relative infectiousness of asymptomatic infections | 0.001 (0.00075, 0.00125) | [26] |
| $b_\mu$ | relative bacterial shedding of asymptomatic individuals | $10^{-7}$ ($7.5 \times 10^{-7}, 1.25 \times 10^{-7}$) | [26] |
| $1/\sigma$ | mean duration of natural immunity | $4 \times 52$ [t] | [26] |
| $r$ | reporting ratio | 0.2 | [26] |
| $k$ | proportion of infections that are symptomatic | 0.2 (0.1, 0.3) | [26] |
| $1/\gamma$ | mean infectious period | 1 (0.5, 1.8) [t] | [26] |
| $V$ | bacteria concentration yielding 50% of catching infection | $10^{-5}$ [cells/mL<sup>3</sup>] | [26] |
| $N$ | total population | 10,911,819 [p] | [26] |
| $p_{\text{under5}}$ | proportion of population under 5 years old | 0.118 | [26] |

## Footnotes

1 http://iridl.ldeo.columbia.edu/SOURCES/.NASA/.GES-DAAC/.TRMM_L3/.TRMM_3B42/.v7/.daily/.precipitation/X/15.0/15.25/RANGEEDGES/Y/12/12.25/RANGEEDGES/T/(01%20Apr%202011)(01%20May%202012)RANGEEDGES/, accessed on March 10, 2019

